# What is the effect of lockdown upon hospitalisation due to COVID-19 amongst patients from a heart failure registry?

**DOI:** 10.1101/2021.02.26.21252336

**Authors:** Hani Essa, Sophia Brousas, Isabel Whybrow Hupptaz, Thomas Salmon, Rajiv Sankaranarayanan

## Abstract

**Introduction:** Coronavirus disease 2019 (COVID-19) is associated with a high risk of mortality especially in patients with cardiovascular conditions such as heart failure. The UK government announced a national lockdown last year to curb the spread of the virus. We conducted this study primarily to ascertain the impact of lockdown upon the incidence of COVID-19 hospitalisation amongst patients with a known diagnosis of heart failure (HF)

**Methods:** This was a retrospective cohort study of 1097 patients from our HF registry who had presented with acute decompensated HF in 2018 and 2019. Incidence and outcomes of hospitalisation due to COVID-19 were analysed in this cohort both during the 1^st^ UK lockdown as well as after the lockdown period. Co-morbidities, frailty index, clinical features, blood results, and heart failure treatments were compared between the 2 groups (COVID versus no-COVID) and between the group of patients who died versus survivors.

**Results:** 50 out of 801 surviving (6.2%) HF patients required hospitalisation due to COVID-19 from March to November 2020; 24 patients (3.1%) during the first lockdown and 26 (3.5%) in the post-lockdown period; p=0.7. In comparison to patients not hospitalised with COVID-19 (“no-COVID group), there was a significantly higher prevalence of co-morbidities amongst HF patients who were hospitalised with COVID-19, such as hypertension (p<0.001), diabetes (p=0.005), ischaemic heart disease (p=0.01) and increased body mass index. 30 day mortality amongst HF patients hospitalised due to COVID-19 was 52%. Rockwood Frailty Score ≥6 (OR 6.530695 % CI:1.8958 to 22.4961; p=0.003) and diabetes (OR 3.82;95% CI 1.13 to 12.95; p=0.03) were independent predictors of 30 day mortality.

**Conclusion:** Our data suggests that the incidence of hospitalisation due to COVID-19 was similar both during as well as post lockdown amongst patients from our HF registry. HF patients with cardiovascular co-morbidities such as obesity, hypertension, diabetes and ischaemic heart disease have a higher risk of hospitalisation due to COVID-19. Diabetes and Rockwood Frailty score are independent predictors of short term mortality. Co-morbidity and frailty scores should be incorporated during initial assessment to help risk-prediction.

## 1.0 Introduction

Coronavirus disease 2019 (COVID-19) is an ongoing international public health crisis caused by severe acute respiratory syndrome coronavirus 2 (SARS-CoV-2) that originated in Wuhan, China at the end of 2019 and rapidly spread globally [1]. COVID-19 has proven to be a multisystem condition with frequent cardiac and respiratory complications. In the United Kingdom (UK) the governmental response to the rapidly accelerating pandemic was to issue a public lockdown from the 23^rd^ of March 2020 and to order a rapid shift in the focus of care in the National Health Service (NHS). The primary objective of a national lockdown was to slow or halt the transmission of the disease in the community particularly in order to protect the most vulnerable population from the critical ill-effects of the disease, until effective treatments or vaccines could be developed. This strategy would also allow healthcare services to be able to effectively manage hospitalisations for all conditions in addition to COVID-19.

Older age, certain cardiovascular risk conditions or risk factors such as ischaemic heart disease (IHD), heart failure (HF), hypertension (HTN), diabetes mellitus (DM), and certain ethnicities (black, Asian and minority ethnicities) have a propensity for association with more severe forms of COVID-19 [2]. There was early recognition that HF patients were potentially more vulnerable and therefore advised to shield at home [3].

There is a relative paucity of data regarding the direct impact of lockdown upon preventing coronavirus transmission and importantly hospitalisation due to COVID-19 amongst patients with chronic conditions such as HF. Whilst there is some published data from the rest of the world [4, 5],as well as the UK [6] regarding the prognosis of HF patients hospitalised with COVID-19 there is not much data regarding the impact of lockdown itself. This study primarily aims to look at the effect of lockdown upon hospitalisations due to COVID-19 amongst patients with HF. In addition, we also ascertained the clinical characteristics and outcomes of HF patients who required hospitalisation due to COVID-19.

## 2.0 Methods

This was a single centre retrospective observational study that was undertaken in a British university hospital looking at the incidence of hospitalisation due to COVID-19 amongst HF patients aged ≥ 18 with COVID-19. We collated co-morbidities, demographics, and mortality from this disease.

This study included patients from our existing database of patients with a confirmed diagnosis of HF. Patients who had presented with acute decompensated HF within the previous two years preceding the onset of the pandemic (i.e, 2018, 2019) were included in this study. Hospitalisations due to COVID-19 amongst this cohort of HF patients, were analysed from electronic hospital records during the period of 1^st^ lockdown in the UK (March to June 2020 “lockdown period”) and compared to COVID-19 hospitalisations post-lockdown (July to November 2020 –”post-lockdown period” i.e, prior to the 2^nd^ lockdown enforced in December 2020. A diagnosis of COVID-19 was considered positive if the patient exhibited typical symptoms along with either a positive SARS-COV polymerase chain reaction (PCR) nasopharyngeal swab result or a typical radiological appearance of COVID-19 pneumonia as reported by a radiologist on a plain film or a CT-thorax scan. We excluded patients with non-classical or indeterminate X-RAY findings.

Demographic characteristics included a review of a patient’s age, sex, ethnicity, body mass index (BMI) and co-existing conditions. We also analysed the prevalence of co-morbidities such as diabetes mellitus, hypertension, ischaemic heart disease (IHD), chronic obstructive pulmonary disease (COPD) or a pre-existing malignant condition and computed the Charlson Co-morbiditiy Index (CCI) [7]. HF clinical risk score (GWTG) and the Rockwood clinical frailty score [7] were also compared between the 2 groups (COVID versus non-COVID). Patient’s clinical information was obtained from outpatient clinical letters, inpatient admission documents, and GP summary care records.

Blood results at the time of admission were also compared between the 2 groups, including haemoglobin, urea, creatinine, serum sodium and NT-proBNP. The incidence of acute kidney injury (AKI) (creatinine rise of 26.5umol/L from baseline or 1.5 to 1.9 X baseline in 48 hours) was also collated [8].

We collected information on the body mass index (BMI) and ethnicities of patients, as both obesity and ethnic background, in particular a background of black, Asian and minority ethnic (BAME) have been linked to higher rates of both COVID-19 and death in the UK for reasons unclear [9].

Medical therapy with prognostic HF therapy such as ACE inhibitors (ACE-I), beta-blockers, mineralocorticoid receptor antagonists (MRA), device therapy was compared between the groups. In addition, we also collected data regarding concurrent immunosuppressant treatment (including chemotherapy, steroids, biologics and immunotherapy) at time of admission, was obtained from admission medication reconciliation.

Descriptive statistics were represented using continuous variables as means with standard deviations (mean ± SD) if normally distributed and compared using a Student’s T test. Non-normal data was described using medians with interquartile ranges and compared using the Mann –Whitney test. Categorical data was expressed as percentages and compared using the chi-square test. The SPSS version 18 software package (SPSS Inc., Chicago, Illinois) was for statistical analysis. Two-sided p values of <0.05 were considered as indicative of statistical significance.

## 3.0 Results

We identified 1097 HF patients from our existing HF registry who had presented with acute decompensated HF in 2018 and 2019. Out of these there were 801 survivors and we retrospectively analysed these patients for episodes of hospitalisation due to COVID-19 both during the 1^st^ period of UK lockdown (March to June 2020) and also post lockdown (July to November 2020). 26 out of 801 HF patients (3%) were hospitalised due to COVID-19 during the 1^st^ period of lockdown and 24 out of the remaining 731 patients (3.1%) (excluding the 1^st^ 26 patients and those who died) sustained hospitalisation due to COVID-19 in the post lockdown period.

Table 1 illustrates the baseline characteristics of patients in our study. HF patients who required hospitalisation due to COVID-19 (COVID group), had a significantly higher prevalence of co-morbidities such as HTN, DM and IHD, in comparison to HF patients who did not require hospitalisation due to COVID (No-COVID group). The COVID group also had a higher BMI (33.67±9.1)) in comparison to the No-Covid group (31.1±8.1; p=0.04). The Charlson Co-morbidity index was also significantly higher in the COVID group (6.5±1.5) in comparison to the no-COVID group (5.7±1; p<0.001). The COVID group was also frailer as estimated by the pre-morbid Rockwood Clinical Frailty Score (COVID group 6.5±1.5 versus 6.1±1.1 in the No-COVID group; p=0.02). Characteristics which were associated with hospitalisation due to COVID are also illustrated in Figure 1.

**Table 1:** Comparison of HF patient characteristics (COVID versus no COVID)

| <u>Baseline Characteristics</u> | <u>COVID (n=50)</u> | <u>NO COVID (n=751)</u> | <u>p</u> |
| --- | --- | --- | --- |
| Age | 75.3 ± 10 | 73 ±14.1 | NS |
| Female | 40% | 46% | 0.4 |
| DM | 27/50 74% | 225/751 30% | <0.001 |
| HTN | 41/50 82% | 466/751 40% | 0.005 |
| IHD | 29/50 58% | 301/751 40% | 0.01 |
| COPD | 11/50 22% | 233/751 31% | 0.18 |
| CKD | 27/50 54% | 370/751 (49%) | 0.52 |
| AF | 25/50 50% | 413/701 | 0.2 |
| Charlson Age adjusted Comorbidity Index | 6.5±1.5 | 6.1±1.1 | 0.01 |
| Rockwood Frailty Index | 5.8±1.9 | 5.1±1.7 | 0.005 |
| Beta blocker | 78% | 653/751 | 0.15 |
| Mineralocorticoid antagonist | 28% | 262/751 | 0.43 |
| ACE/ARB/ARNI | 65% | 81% | 0.05 |
| Device therapy | 7% | 60/751 | 0.95 |
| Average length of stay | 15.6 (± 14.8)<br>Median 14.5 (3-57) | 8.6<br>8 (1-43) | <0.01 |
| HFpEF | 36% | 32% | 0.92 |
| BMI | 33.67 (±9.1) | 31.1±8.1 | 0.04 |
| NTpro-BNP | 6242 (415-24000) | 3564 (515-7000) | 0.07 |
| Hb | 118.5± 19.2 | 119±20.5 | 0.83 |
| Urea | 6.9± 3.9 | 8±4 | 0.2 |
| Creatinine | 154±71 | 128±69 | 0.05 |
| Sodium | 136±5.3 | 139±4.5 | <0.001 |
| GWTG | 43±7.1 | 38.8±6.4 | <0.001 |
| Table 1. Demographic data and background of patients admitted with COVID vs Non-COVID. P<0.05 is taken to mean statistical significance. |  |  |  |

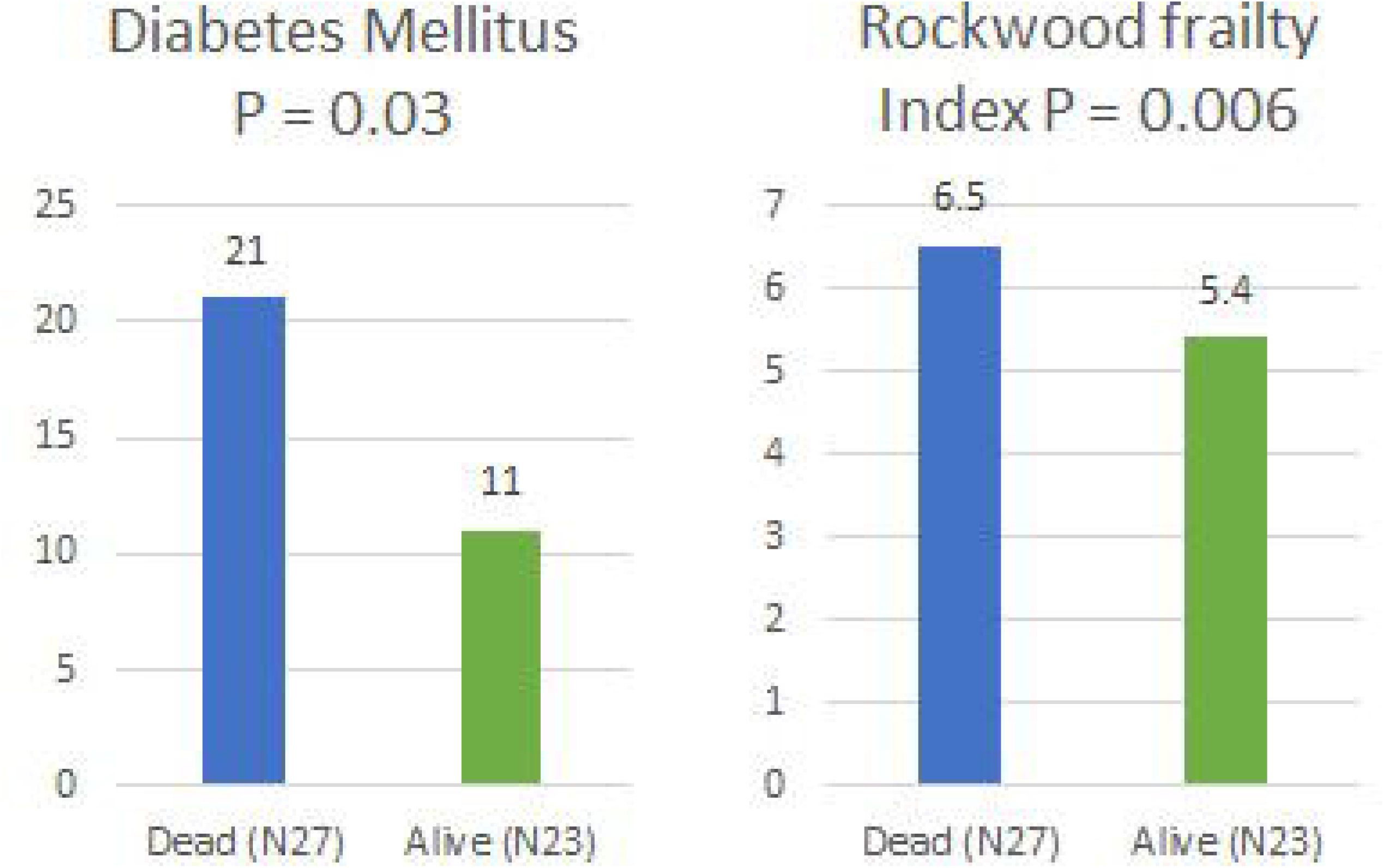

The proportion of patients with impaired LV systolic function (HFrEF and HFmrEF) was comparable between the 2 groups as were the baseline prognostic medications (beta-blockers, ACE inhibitors and mineralocorticoid receptor antagonists).

Table 2 compares the characteristics between HF patients who died within 30 days of hospitalisation due to COVID-19 and the survivors. Whilst the incidence of diabetes hypertension and frailty were significantly higher amongst the group that died within 30 days, multivariate regression analysis demonstrated that only diabetes (OR 3.82;95% CI 1.13 to 12.95; p=0.03) and Rockwood Frailty Score (≥6; OR 6.530695 % CI:1.8958 to 22.4961; p=0.003) were independent predictors of mortality (Figure 2). HF patients hospitalised with COVID also had a longer hospital stay than for HF (median 14.5 days vs. 8 days; p<0.001).

**Table 2:** Comparison of HF patient characteristics (DEAD versus no ALIVE)

| Baseline Characteristics | DEAD<br>(n=27) | ALIVE (23) | p |
| --- | --- | --- | --- |
| Age | 77.8±8.9 | 73.9 ±10.1 | 0.2 |
| Female |  |  |  |
| DM | 21/6 | 11/12 | 0.03 (OR 3.82;95% CI 1.13 to 12.95) |
| HTN | 21/27 | 12/23 | 0.06 OR 3.2<br>(95% CI 0.9456 to 10.8858) |
| IHD | 15/12 | 10/13 | 0.4 (OR 1.63 CI 0.5 TO 5) |
| COPD | 7/27 | 4/23 | 0.5 (OR 0.9 CI 0.3 TO 2.96) |
| CKD | 15/27 | 12/23 | 0.81 (OR 1.15 CI 0.38 to 3.5) |
| AF | 14/27 | 11/23 | 0.37 |
| Charlson Age adjusted Comorbidity Index | 6.5 ± 1.6 | 5.9 ±1.3 | 0.15 |
| Rockwood Frailty Index | 6.2±1 | 5.4±1 | 0.006<br>(≥6; OR 6.530695 % CI:1.8958 to 22.4961; p=0.003) |
| Beta blocker | 71% | 80% | 0.22 |
| Mineralocorticoid antagonist | 31% | 24% | 0.45 |
| ACE inhibitor | 61% | 71% | 0.18 |
| HFrEF |  |  | 0.9 |
| BMI | 33.6±8 | 31.8±7.5 | 0.4 |
| NTpro-BNP | 6807(1810-24305) | 5700 (1653-17000) | 0.91 |
| Hb | 121± 20 | 115± 18 | 0.33 |
| Lymphocyte count | 1±0.4 | 1.1±0.5 | 0.5 |
| BUN | 6.9±4 | 6.7±3.5 | 0.9 |
| Creatinine | 153± 98 | 150± 65 | 0.9 |
| Sodium | 134.3± 5.9 | 136.7± 4.3 | 0.42 |
| BP | 134±29 | 146±32 | 0.17 |
| HR | 88±20 | 78±19 | 0.2 |

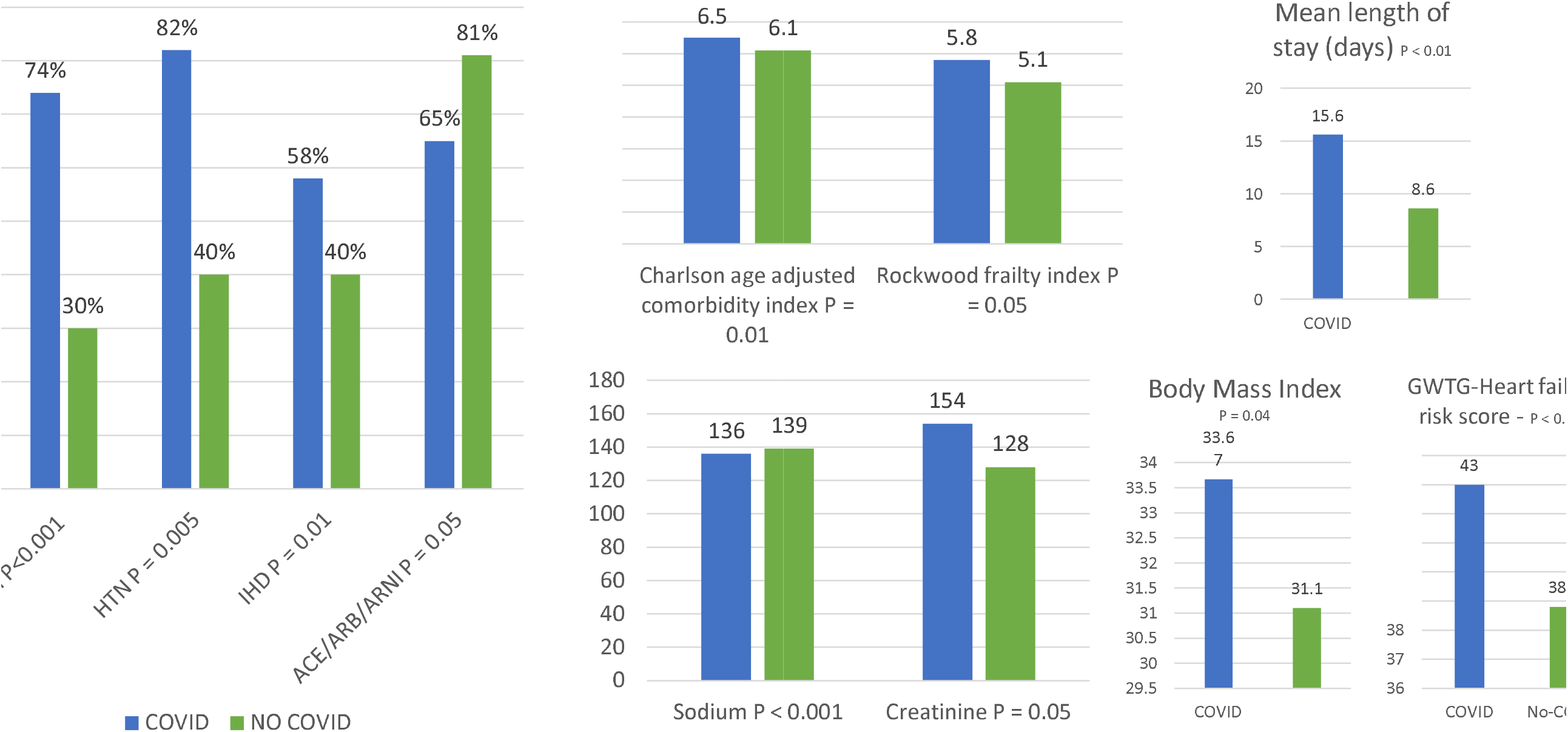

## 4.0 Discussion

Our study has shown a similar incidence of hospitalisation due to COVID-19 pre- and post-lockdown amongst patients previously hospitalised with acute decompensated HF within the two years preceding the pandemic. The overall incidence of hospitalisation due to COVID-19 in our HF cohort (3.1%) during these respective periods of study, was similar to the incidence of hospitalisation due to COVID-19 during the first wave in England (3.5%) [10]. It can only be surmised that patients with HF have been taking adequate shielding precautions in view of media reports of higher complications due to COVID-19 amongst patients with cardiovascular co-morbidities. As we included HF patients with hospitalisation due to decompensated HF during the previous two years, it is possible that this cohort of patients took shielding precautions especially seriously. It is also possible that the anxiety felt by HF patients and their reluctance to attend hospital may also have resulted in reduced hospitalisations due to COVID-19 [11].

We have shown that in patients with a known diagnosis of HF, 30-day mortality due to COVID-19 hospitalisation was high (54%). This is comparable to results from 2 other UK studies which also demonstrated an approximate doubling of in-hospital and 30-day mortality following hospitalisation due to COVID-19 in HF patients [6, 12]. Furthermore, these patients suffer from prolonged length of stay and over a third are complicated by AKI. The existing literature broadly agrees with our data demonstrating that HF patients hospitalised due to COVID-19 have a significantly higher BMI and also a high co-morbidity burden as evidenced by the higher Charlson co-morbidity Index and these particularly included cardiovascular co-morbidities such as hypertension, IHD and diabetes. However, diabetes and frailty (particularly clinical frailty scale of ≥6) were independent predictors of short-term mortality. A recently published study of a much larger cohort of HF patients hospitalised with COVID-19 also demonstrated that diabetes is an independent risk factor for mortality [13]. It is therefore important that the Charlson Co-morbiditiy index as well as Rockwood Clinical Frailty Scale are assessed during initial clinical risk assessment as risk-prediction tools.

Several limitations must be acknowledged when interpreting this data. First, this is a retrospective observational study dependent on accurate electronic records with the inherent limitations of this type of design. Second, whilst the study looked at 801 patients in total, the actual cohort with COVID-19 hospitalisation consisted of 50 patients. Whilst it is difficult to draw firm conclusions from this small cohort, several findings are in line with other studies. Third, the data utilised is from a single centre and it is difficult to extrapolate outcomes across the UK to other patient groups. Finally, since diagnosis was based on a swab or radiological findings it is possible that patients with false negative tests were missed from the analysis.

## 5.0 Conclusion

Our study did not show a significant difference in hospitalisation of HF patients due to COVID-19 pre- and post-lockdown in the UK. HF patients have a high co-morbidity burden that possibly predisposes them to complications of COVID-19 such as hospitalisation but also portends higher mortality particularly in presence of diabetes and higher frailty. The Charlson co-morbidity score and Rockwood Clinical Frailty Score should be incorporated as a part of routine clinical assessment in these patients to aid mortality risk-prediction.

## Data Availability

Data available on request

## Abbreviations

HF: Heart failure
HFrEF: Heart failure with reduced ejection fraction
HFpEF: heart failure with preserved ejection fraction
DM: Diabetes mellitus
HTN: hypertension
IHD: Ischemic heart disease

## Notes

### Competing Interest Statement

The authors have declared no competing interest.

### Funding Statement

Nil funding

### Author Declarations

Local Aintree university hospital ethics committee.

## References

[1] Viruses CSGotICoTo. The species Severe acute respiratory syndrome-related coronavirus: classifying 2019-nCoV and naming it SARS-CoV-2. Nat Microbiol. 2020;5:536–44.

[2] Zhou F, Yu T, Du R, Fan G, Liu Y, Liu Z, et al. Clinical course and risk factors for mortality of adult inpatients with COVID-19 in Wuhan, China: a retrospective cohort study. Lancet. 2020;395:1054–62.

[3] Powis S. 200321_COVID-19_CMO_MD_letter-to-GPs_FINAL_2.pdf. 2020.

[4] Inciardi RM, Adamo M, Lupi L, Cani DS, Di Pasquale M, Tomasoni D, et al. Characteristics and outcomes of patients hospitalized for COVID-19 and cardiac disease in Northern Italy. Eur Heart J. 2020;41:1821–9.

[5] Alvarez-Garcia J, Lee S, Gupta A, Cagliostro M, Joshi AA, Rivas-Lasarte M, et al. Prognostic Impact of Prior Heart Failure in Patients Hospitalized With COVID-19. J Am Coll Cardiol. 2020;76:2334–48.

[6] Chatrath N, Kaza N, Pabari PA, Fox K, Mayet J, Barton C, et al. The effect of concomitant COVID-19 infection on outcomes in patients hospitalized with heart failure. ESC Heart Fail. 2020.

[7] Charlson ME, Pompei P, Ales KL, MacKenzie CR. A new method of classifying prognostic comorbidity in longitudinal studies: development and validation. J Chronic Dis. 1987;40:373–83.

[8] Section 2: AKI Definition. Kidney Int Suppl (2011). 2012;2:19–36.

[9] Kirby T. Evidence mounts on the disproportionate effect of COVID-19 on ethnic minorities. Lancet Respir Med. 2020;8:547–8.

[10] Report 41 - The 2020 SARS-CoV-2 epidemic in England: key epidemiological drivers and impact of interventions. @imperialcollege; 2021.

[11] Sankaranarayanan R, Hartshorne-Evans N, Redmond-Lyon S, Wilson J, Essa H, Gray A, et al. The impact of COVID-19 on the management of heart failure: a United Kingdom patient questionnaire study. ESC Heart Fail. 2021.

[12] Doolub G, Wong C, Hewitson L, Mohamed A, Todd F, Gogola L, et al. Impact of COVID-19 on inpatient referral of acute heart failure: a single-centre experience from the south-west of the UK. ESC Heart Fail. 2021.

[13] Bhatt AS, Jering KS, Vaduganathan M, Claggett BL, Cunningham JW, Rosenthal N, et al. Clinical Outcomes in Patients With Heart Failure Hospitalized With COVID-19. JACC Heart Fail. 2021;9:65–73.

